# The impact of the COVID-19 pandemic on secondary prevention of Rheumatic Heart Disease in Western Uganda

**DOI:** 10.1101/2025.03.14.25323941

**Authors:** Xinpeng Xu, Emily Chu, Jenifer Atala, Rosemary Kansiime, Miriam Nakitto, Emma Ndagire, Haddy Nalubwama, Emmy Okello, Jafesi Pulle, David A. Watkins, Chris T. Longenecker, Gloria D Coronado, Yanfang Su

## Abstract

**Background:** Regular outpatient visits to healthcare facilities for benzathine penicillin injections are the primary means of secondary prevention for patients with Rheumatic Heart Disease (RHD). It has been widely demonstrated that individual access to healthcare has been disrupted by the COVID-19 outbreak, but its impact on RHD patients has not been studied.

**Methods:** Using a stratified random sampling strategy, we conducted a baseline and follow-up survey of RHD patients in Western Uganda between November 2019 and February 2020, and November 2020 and February 2021, and obtained 486 month-patient outpatient records. A fixed effect model and event-study analysis method were employed to investigate the impact of the COVID-19 pandemic on RHD patients’ secondary prevention.

**Results:** Prior to the outbreak of COVID-19, the average number of secondary prevention visits for RHD patients in Uganda were 9.48 per patient per year, which was far below the criteria of clinical guidelines. The secondary prevention visits decreased by an average of 0.12 visits per month in the COVID-19 pandemic, but the decrease was not statistically significant (*P*>0.05). The number of monthly secondary prevention visits among patients with RHD exhibited a gradual upward trend over time. The number of secondary prevention visits increased significantly (*P*<0.05) in the eight months following the COVID-19 outbreak in western Uganda.

**Conclusions:** RHD patients in Western Uganda failed to adhere to the recommended frequency of secondary antibiotic prophylaxis prior to the outbreak of the COVID-19 pandemic, which decreased further during the pandemic. Improving the accessibility and convenience of treatment for RHD patients is a crucial step in enhancing the frequency of secondary prevention. Future pandemic preparations should consider the demands of secondary prevention among RHD patients.

## Introduction

Rheumatic Heart Disease (RHD) is a disease of poverty which impacts approximately 41 million people globally [1] and is most prevalent in socioeconomically disadvantaged countries[2]. Pooled studies reveal that the prevalence of RHD is 26.1% and 11.3%, based on the World Heart Federation and World Health Organization criteria respectively[3]. RHD develops from Acute Rheumatic Fever (ARF), an inflammatory, abnormal[4] autoimmune response caused by an untreated Group A Streptococcus infection. This results in a progressive, irreversible damage to cardiac valves and often fatal complications, most notably heart failure, atrial. fibrillation and stroke. To prevent the recurrence of ARF and the progression of RHD[5], secondary antibiotic prophylaxis (SAP), typically in the form of Benzathine penicillin G (BPG), is often administered to RHD patients in outpatient visits every three to four weeks. BPG has been shown to reduce the recurrence of ARF and progression of RHD with optimum adherence, defined as receiving ≥80% of prescribed injections. Given the frequency of interactions of RHD patients with the healthcare system, a lapse of access to healthcare can be detrimental to RHD patients[2]. Recently, the coronavirus disease (COVID-19) pandemic has disrupted access to care for many patients, risking the progression of RHD in RHD patients and other adverse outcomes[2]. To better prepare for future pandemics and ensure the consistent provision of essential healthcare services to RHD patients in Uganda despite unexpected circumstances, it is crucial to understand the extent to which RHD outpatient visits have been interrupted by the COVID-19 pandemic.

Officially declared as a global pandemic of concern by the World Health Organization (WHO) in March of 2020, the COVID-19 pandemic resulted in lockdowns and social distancing orders, among other policies, around the world[6]. These measures were designed to mitigate the risk of spreading COVID-19 virus but as a result, have severely disrupted essential health services. Globally, the COVID-19 pandemic has significantly impacted healthcare utilization, with a median 37.2% reduction in healthcare utilization between pre-pandemic and pandemic periods[7]. In a large integrated healthcare system located in the United States, inpatient, emergency department, and outpatient service utilization were reduced significantly during the pandemic, by margins of 30.2%, 37.0%, and 80.9% respectively[8]. This global reduction in healthcare utilization was particularly drastic for people living with non-communicable diseases, with many patients requiring treatment that could not be administered with alternate healthcare methods during the pandemic, such as telemedicine[9]. According to a survey taken by WHO in 2020, the pandemic partially or completely disrupted hypertension treatment in 53% of the countries surveyed, cancer treatment in 42%, and diabetes treatment in 49%[10]. Internationally, cardiovascular practices and procedure volumes decreased 64% from March 2019 to April 2020, with acute cardiovascular disease admissions decreasing up to 50%[11,12].

In low- and middle-income countries (LMICs)[13,14], such as Uganda, where less economically advantaged conditions often result in a greater disruption of medical supplies and care[14], the COVID-19 pandemic has resulted in even worse healthcare utilization rates. More than 53% of LMICs reported partially or completely impaired services for non-communicable diseases (NCDs) and related complications, with only 42% of LMICs attempting to focus on NCD services during the COVID-19 pandemic as opposed to 72% of high-income countries[14]. As a LMIC, healthcare utilization in Uganda has been greatly reduced. Resulting from the pandemic, the decline in the number of outpatient consultations, a proxy for general service utilization, is estimated to be between 10% and 20% in Uganda[15]. Facility deliveries in March 2020 as compared to January 2020 were reduced by 29%, and the volume of initiations of isoniazid-preventative therapy to prevent secondary tuberculosis was reduced by 75%[16]. Certain healthcare outcomes worsened as well in Uganda due to service disruptions during the COVID-19 pandemic, with child mortality was estimated to have increased by at least 5%[15].

In particular, many RHD patients in Uganda, who require consistent receipt of secondary prophylaxis and must frequently interact with the healthcare system, are also especially prone to encountering interruptions in access to care[2]. To our knowledge, no studies have studied the impact of the COVID-19 pandemic on RHD patients’ secondary prevention. This study aims to investigate the impact of the COVID-19 pandemic on the secondary prevention by RHD patients in Uganda and to provide a useful reference for the treatment of RHD patients during the pandemic.

## Methods

### Data source

The data used in this study were from a longitudinal survey conducted in western Uganda[17]. We recruited RHD patients from the Uganda National RHD Registry using a multi-stage random sampling procedure. Firstly, based on previous RHD-related research cooperation and infrastructure, we randomly selected a referral hospital in the western region and intended to recruit one-third of patients over the age of 10 who registered in the RHD registry and received treatment. However, due to the outbreak of the COVID-19 pandemic, the ethics review committees of the United States and Uganda in this study halted nearly all research activities after March 2020. We surveyed 21 RHD patients in Western Uganda for the baseline survey. In July 2020, when the ethical review board authorized the study to proceed, a follow-up survey of RHD patients was conducted. The surveys involved the household and individual characteristics of RHD patients and the outpatient records of each patient in the last 12 months, which allowed us to analyze the impact of the COVID-19 pandemic on outpatient healthcare utilization of RHD patients. Figure 1 illustrates the period during which baseline and follow-up surveys were conducted in Western Uganda.

**Figure1.**
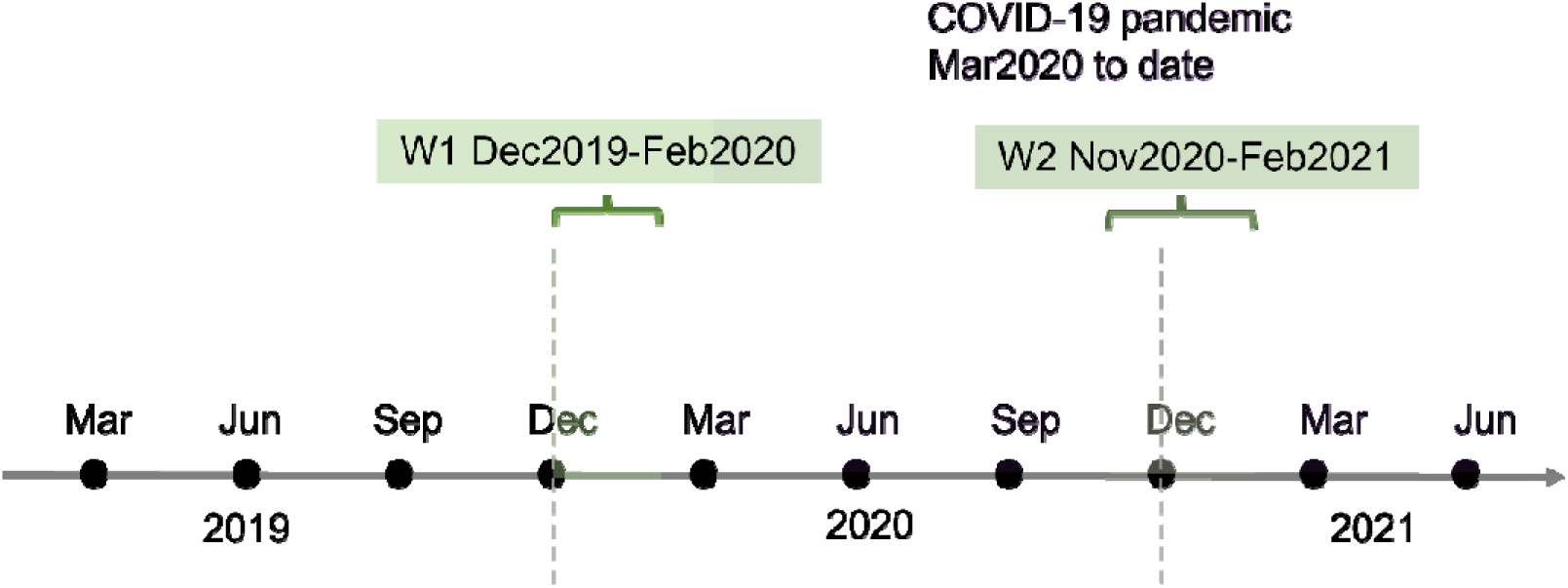
Surveys Taken in Western Uganda. Note: In Figure1, W1 and W2 are baseline and follow-up surveys conducted in the western Uganda, respectively.

The survey time varied for each RHD patient, and the number of outpatient visits utilized by RHD patients was recorded between December 2018 and February 2021. From December 2018 through February 2021, we compiled a balanced panel data that tallied the number of outpatient visits per month for each individual. If a respondent’s survey period did not encompass a particular month, the number of outpatient visits in that month was coded as missing (For instance, if a patient has a baseline survey time of February 2020 and recalls 12-month visits from March 2019 to February 2020, the patient’s visits in December 2018, January 2019, and February 2019 are coded as missing values). Three individuals in the data lacked outpatient healthcare utilization information, so our final sample for analysis consisted of 486 observations from 18 RHD patients (27 months for each RHD patient).

### Variables

#### Healthcare utilization

We used the number of self-reported outpatient visits as a proxy for healthcare utilization among RHD patients. Each patient was asked about each outpatient visit record in the past 12 months. We added the number of visits per patient per month and utilized the monthly outpatient visits as the dependent variable.

#### COVID-19 pandemic

The first COVID-19 case was reported in Uganda on March 22, 2020. Hence, we selected March 2020 as the timepoint of the start of pandemic[18]. We created a dummy variable that represents during the COVID-19 pandemic (after March 2020) when its value is 1 and prior to the pandemic (before March 2020) when its value is 0.

#### Covariates

In addition to the COVID-19 pandemic, we incorporated other variables affecting the healthcare utilization of RHD patients in the model, including the patients’ age, gender, education level, monthly household income per capita, and EQ-5D index. Age is a continuous variable that indicates the true age of the patient. Gender is a dummy variable, with the value 1 representing female and 0 representing male. Education level is a dichotomous variable, with a value of 1 indicating that the patient has completed secondary school education or higher, and a value of 0 indicating that the patient has completed a primary school education or less. Monthly household income per capita is a continuous measure, determined by factoring in inflation and the currency conversion, and eventually expressed in 2020 US dollars. In our study, we measured the health status of RHD patients using the EQ-5D-3L. EQ-5D-3L is a commonly used generic scale for measuring individual health status[19]. The EQ-5D-3L consists of five dimensions, namely mobility, self-care, usual activities, pain/discomfort, and anxiety/depression, with three alternative responses for each dimension: 1 = no problems, 2 = some/moderate problems, and 3 = extreme problems. We utilized the Zimbabwe EQ-5D-3L value set determined by the time trade-off method to convert each unique choice combination to a single index for each RHD patient[20].

### Statistical analyses

#### Descriptive statistics

Descriptive statistics were used initially to analyze the characteristics of the study population in the baseline and follow-up survey. Mean and standard deviation are two descriptive statistics used for continuous variables. Frequency and percentage calculations are used for descriptive statistics when dealing with categorical data.

In accordance with the purpose of our study, we calculated the average number of outpatient visits per patient per month by dividing monthly outpatient visits by the number of patients per month. This allowed us to clearly demonstrate changes in the number of outpatient visits of RHD patients during different time periods. We plotted the outpatient visits per patient per month of RHD patients to observe the differences among each period, as well as the changes prior to and following the outbreak of the COVID-19 pandemic.

#### Fixed effect model

We utilized a fixed effect model to examine the impact of the COVID-19 pandemic on healthcare utilization among RHD patients. The model is set as follows:

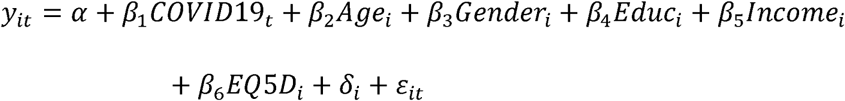

Where *i*(*i* = 1, ·,21) denotes the patient with RHD and *t* denotes the month. The dependent variable *y_it_* represents the number of outpatient visits of the *i_th_* RHD patient in month *t*. The individual-specific fixed effects *δ_i_* controls unobservable individual heterogeneity that does not change over time. *β*_1_ is the estimator that this study focuses on, which represents the changes in outpatient healthcare utilization following the COVID-19 pandemic relative to prior periods. A positive *β*_1_ suggests an increase in RHD patients’ outpatient visits following the COVID-19 pandemic. When *β*_1_ is negative, however, it indicates that the monthly number of outpatient visits for RHD patients has dropped since the COVID-19 pandemic.

#### Event study

Using the event study method, we examined the change in outpatient visits of RHD patients before and after the outbreak of COVID-19 pandemic, with reference to previous research[21]. The special model is set as follows:

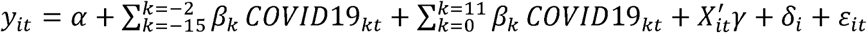

Where *y_it_* represents the number of outpatient visits of individual *i* in month *t*, and COVID19*_kt_* represents the indicator between month *t* and the month of the COVID-19 outbreak. To demonstrate the impact of the COVID-19 pandemic, the model excludes the month preceding the event (*t*=-1). *X_it_* represents the other covariates in Equation (1), including age, sex, education, income, and EQ-5D. *δ_i_* represents the individual fixed effect. All analyses were conducted using Stata 16.0 statistical software (Stata, College Station, Texas, USA).

## Results

### Baseline Characteristics of respondents

Table 1 showed the basic characteristics of the 18 RHD patients at the baseline and follow-up surveys. The average age of RHD patients was 28.22 ± 15.55 and 29.39 ± 15.49 years old at baseline and follow-up respectively. 61.11% of RHD patients were female. 50.00% of RHD patients had completed at least secondary schooling at baseline, while 66.67% had at follow-up. At baseline and follow-up respectively, the average monthly household income per capita of RHD patients in our sample was 37.35 and 21.57 USD, and the average EQ-5D score of RHD patients was 0.78 and 0.79. The average monthly outpatient visits for RHD patients were 0.79 at baseline and 0.63 at follow-up.

**Table 1.** Summary Statistics of baseline characteristics among RHD patients (n=18)

| Variables | Baseline | Follow-up |
| --- | --- | --- |
| Age, Mean (S.D.) | 28.22 (15.55) | 29.39 (15.49) |
| Female, n (%) | 11 (61.11%) | 11 (61.11%) |
| Secondary or above education, n (%) | 9 (50.00%) | 12 (66.67) |
| Monthly Household Income per Person (USD), Mean (S.D.) | 37.35 (51.93) | 21.57 (17.26) |
| EQ-5D, Mean (S.D.) | 0.78 (0.14) | 0.79 (0.09) |
| Outpatient Visits per patient per month, Mean (S.D.) | 0.79 (0.98) | 0.63 (0.59) |

### Outpatient visits per patient per month

We summed up the number of outpatient visits per patient per month based on the outpatient records of RHD patients from the past 12 months. If a patient did not self-report an outpatient record in a given month during the past year, the number of outpatient visits was coded as 0. Figure 2 depicts the outpatient visits per patient per month based on a 12-month recall period.

**Figure 2.**
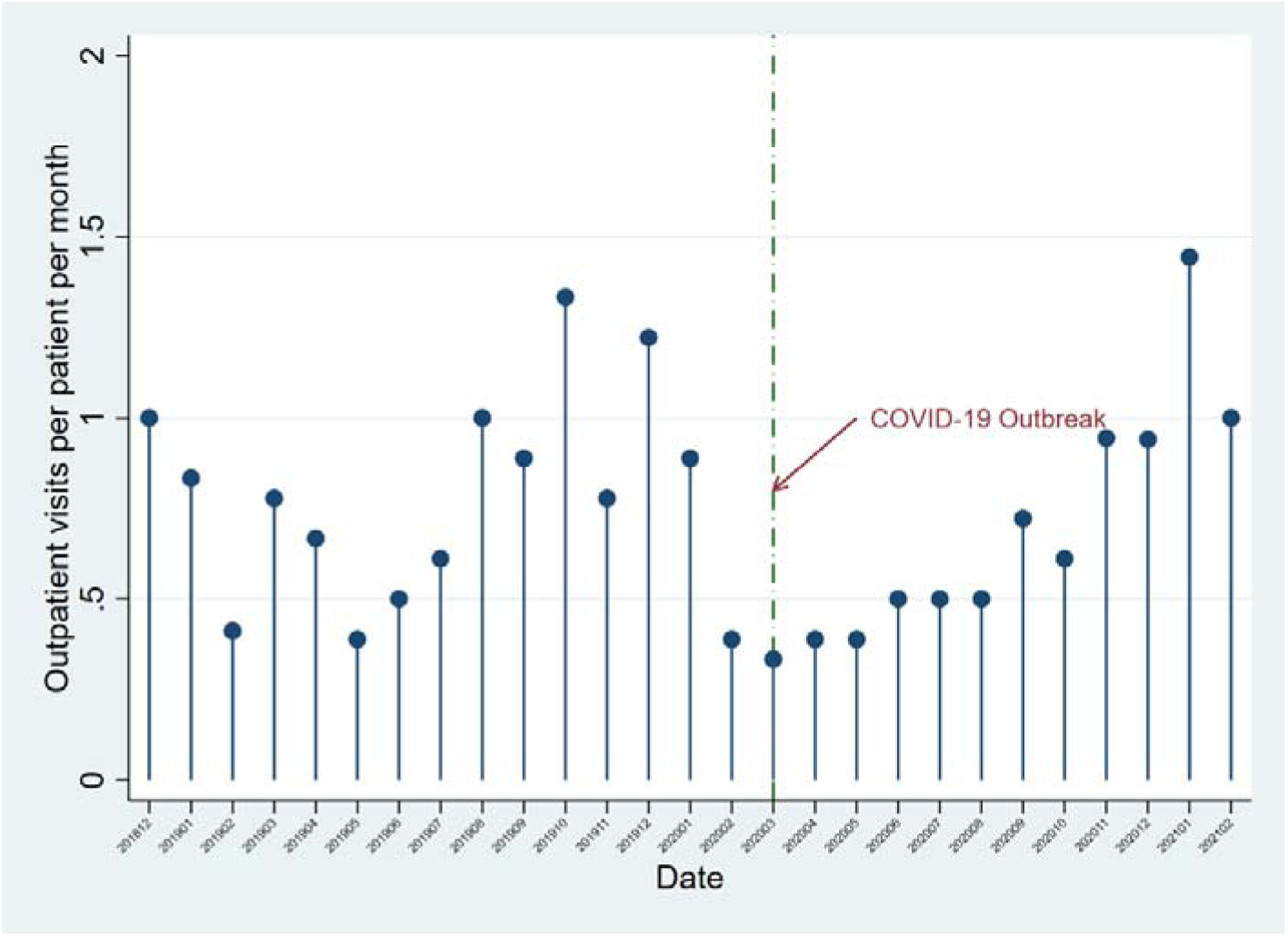
Outpatient Visits per patient per month.

In Figure 2, the number of outpatient visits per patient per month was fewer than 1.5 across all 12 months. Additionally, the number of outpatient visits per month fluctuated significantly month to month.

### The impact of COVID-19 pandemic on healthcare utilization among RHD patients

We used the fixed effect model to estimate the changes in the number of monthly outpatient visits of RHD patients before and after the outbreak of the COVID-19 pandemic by adding various covariates to the model (Table 2). After COVID-19 outbreak in Uganda, the average number of monthly outpatient visits for RHD patients increased by 0.25 without the addition of covariates, which is not statistically significant. However, after adding age, gender, and education to the model, the average number of visits per month for RHD patients decreased by 0.016, although the difference was not statistically significant. After further controlling for RHD patients’ monthly household income per capita and EQ-5D, the estimates results indicated a mean decrease of 0.12 visits per month for RHD patients after the COVID-19 outbreak. This effect was not considerable and was not statistically significant (P>0.05).

**Table 2.** The impact of COVID-19 pandemic on the monthly outpatient visits of RHD patients: Fixed effect model.

|  | Model1 | Model2 | Model3 |
| --- | --- | --- | --- |
| Variables | Outpatient Visits in the<br>Month | Outpatient Visits in the<br>Month | Outpatient Visits in the<br>Month |
| COVID-19 outbreak (Ref: Before) | 0.25<br>(0.54) | -0.02<br>(0.56) | -0.12<br>(0.51) |
| Age (Ref: Mean) |  | 0.19*<br>(0.10) | 0.18**<br>(0.08) |
| Female (Ref: Male) |  | - | - |
| Secondary or above education (Ref:<br>Primary or less) |  | 0.28<br>(0.19) | 0.41**<br>(0.19) |
| Monthly Household Income per Person<br>(USD) |  |  | -0.003**<br>0.001 |
| EQ-5D |  |  | 0.54<br>(0.60) |
| Year-month fixed effects | Control | Control | Control |
| Observations | 438 | 438 | 438 |
| Adjusted R-squared | 0.10 | 0.11 | 0.12 |

Figure 3 depicted the change in outpatient service utilization by RHD patients before and after the COVID-19 pandemic outbreak. The number of RHD visits was significantly higher in the six months preceding the outbreak. Although the number of outpatient visits for RHD patients decreased immediately after the COVID-19 outbreak, this decline was not statistically significant. As time went on, the number of outpatient visits by RHD patients increased progressively. Eight months after the COVID-19 outbreak in western Uganda, the number of RHD outpatient visits increased significantly.

**Figure 3.**
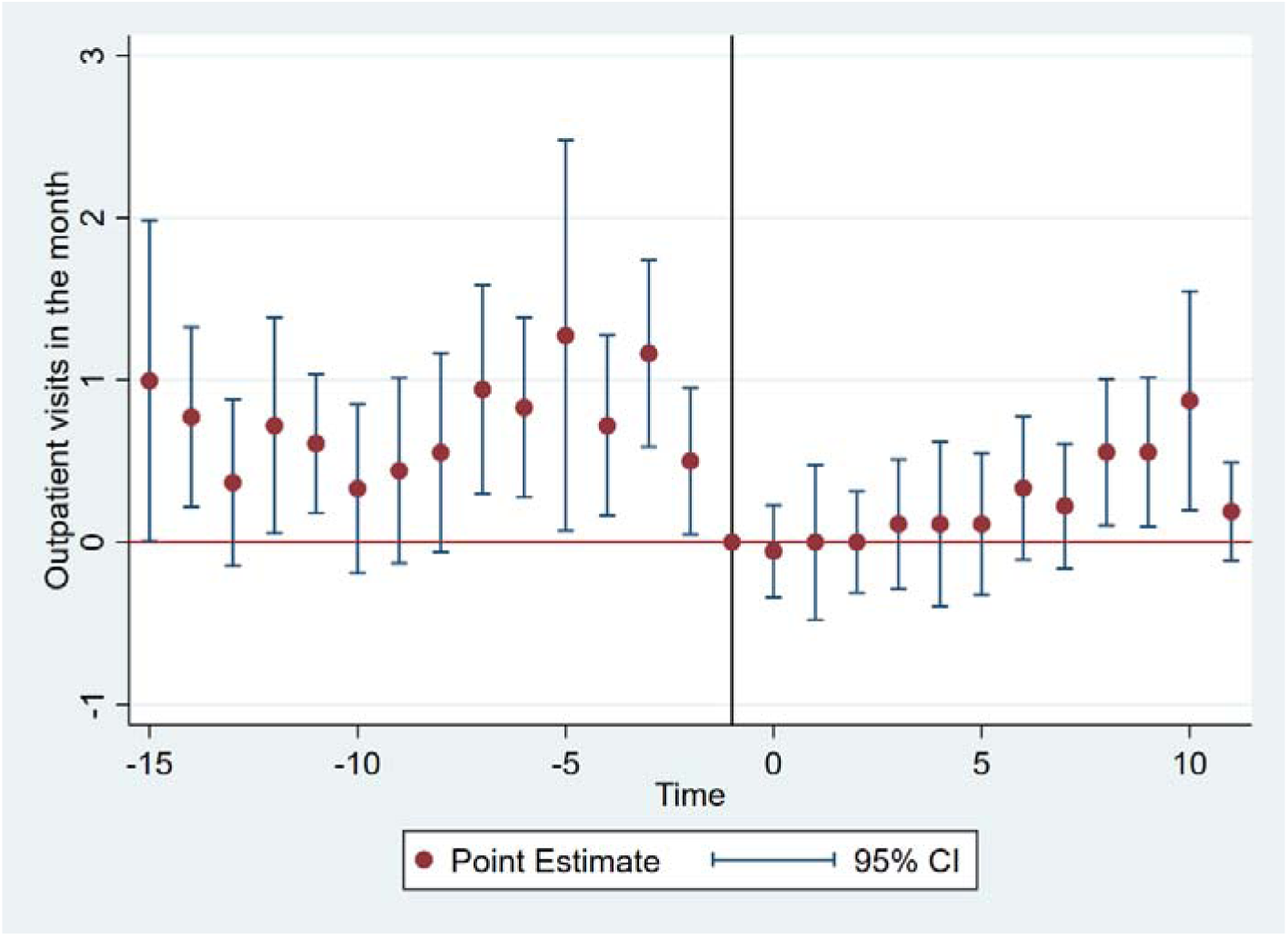
Event study analysis results

## Discussion

This study employed longitudinal survey data collected before and during the COVID-19 pandemic in Western Uganda to estimate the impact of COVID-19 on outpatient healthcare utilization among RHD patients. The results demonstrated that the level of outpatient service utilization for RHD patients was inadequate even before the pandemic, and that the level of outpatient service utilization declined even further during the pandemic. The fixed effect model and event study method consistently demonstrated that the monthly outpatient service utilization of RHD patients after the COVID-19 pandemic decreased immediately compared to periods preceding the pandemic. However, this effect was not significant. After the COVID-19 pandemic, the number of outpatient visits for RHD patients has gradually increased over time.

Our findings demonstrates that RHD patients suffer a immediate decline in outpatient healthcare utilization following the outbreak of the COVID-19 pandemic, which is consistent with the findings of other studies investigating the impact of COVID-19 on healthcare utilization[7,22–27]. Several factors may have contributed to this result. Primarily, the allocation of health resources in Uganda could have limited adequate provision of healthcare services. During the COVID-19 pandemic, many medical resources in Uganda may have been committed to the diagnosis and containment of the epidemic, resulting in fewer resources available for diseases other than COVID-19[24]. Similar cases have been reported in other nations[28]. The pandemic has also presented significant obstacles for patient action. Concerns about the risk of COVID-19 infection at the time of treatment may have caused patients to delay or forego medical treatment[9,25]. Also, the policies adopted by countries or regions to prevent the spread of the epidemic may have also reduced the likelihood of patients’ healthcare utilization[9,29], such as the social distancing policy and road blocks and disruptions in the supply chain of medicine. Overall, delays in diagnosis and secondary prevention are likely to exacerbate the deterioration of RHD patients, resulting in negative long-term consequences[30].

Our study also revealed that the number of monthly outpatient visits for RHD patients in Western Uganda was below the required amount even prior to the COVID-19 pandemic. Studies have demonstrated that secondary prophylaxis with penicillin is the most cost-effective RHD prevention method[31]. The effect of intramuscular penicillin injection is superior to oral penicillin, and 2-3 weeks of benzathine penicillin G injection for secondary prophylaxis is more effective than 4 weeks[32,33]. However, our findings illustrate that RHD patients in Western Uganda did not meet the recommended frequency of secondary prevention before the COVID-19 pandemic. During the pandemic, the injection frequency of secondary prevention was further reduced. There is still significant room for improvement in providing adequate secondary prevention to RHD patients in Uganda even outside of the context of the COVID-19 pandemic.

This study has relevant policy implications. In light of the negative effects of the COVID-19 pandemic on the secondary prevention of RHD patients, it is necessary to take appropriate measures. Improving the ability of nursing personnel in community health centers to deliver benzathine penicillin injections to RHD patients could minimize the risk of COVID-19 infection in RHD patients and increase the accessibility of their prevention and treatment in the short term. Long-term research and development of oral antibiotics for RHD patients should be strengthened to increase their convenience and compliance. In addition, due to recall bias in self-reported healthcare utilization, it is recommended to shorten the recall period to optimally less than 6 months to control measurement error when conducting retrospective surveys of patients. In this study, an attempt was made to incorporate data from administrative or health faculty records to supplement patient-reported data, such as with data from the Ugandan Rheumatic Heart Disease registry. However, due to the lower data quality and incomplete nature of many of these records, especially after the COVID-19 outbreak, we chose to only use self-reported data. For future studies, it is recommended to link patient-level sociodemographic surveys with administrative or health facility health records to acquire accurate and reliable data when possible.

## Limitations

This study has some limitations. First, the sample size used in this study is small, possibly causing other variables in our regression to not be significant; we anticipate significant results when the sample size is increased. Second, as mentioned in our study, by shortening the recall period, we hope to be able to control recall bias-induced underestimation of healthcare utilization by RHD patients. In addition, we suggested in our survey that patients could exhibit receipts or invoices related to their clinic expenses to confirm the records’ accuracy as much as possible; however, the issue of self-reported potential recall bias is still difficult to overcome. Existing studies indicate that using data from databases in medical institutions or administrative departments is the gold standard for patient information[34], but that institutions and departments also have quality and comprehensive issues[35]. In the future, it would be preferable to examine the effect of the COVID-19 pandemic on the secondary prevention of RHD in RHD patients by combining Uganda’s national RHD registry data and retrospective survey data for RHD patients.

## Conclusion

Our results showed that the number of monthly outpatient visits for RHD patients in Western Uganda was below the required amount even prior to the COVID-19 pandemic, and secondary prevention for RHD patients after the COVID-19 pandemic has further decreased. Pandemic preparedness for public health emergencies similar to the COVID-19 pandemic will need to take the need of RHD patients for secondary prevention into consideration. Improving the convenience of penicillin injections in the short term and developing oral antibiotics in the long term are realistic ways to increase the use of secondary prophylaxis for RHD patients and mitigate the harmful impacts of the pandemic on their health. In retrospective studies of routine healthcare, it is necessary to be aware of potential recall bias, which may significantly impact the accuracy of data. To mediate these possible inaccuracies, it is recommended to decrease the self-reported recall window of patients or integrate retrospective surveys with data from health facilities or administrative organizations. Integration of administration or health faculty records with self-reported data can be made more feasible in developing countries by strengthening health information systems, especially during pandemic periods.

## Data Availability

The raw data supporting the conclusions of this study are available to authors upon reasonable request.

## Ethical statement and consent to participate

This study was approved by the Makerere University School of Medicine Research and Ethics Committee (REC RF 2018-082) and by the Uganda National Council for Science and Technology (SS 5081). Written informed consent to participate in this study was provided by the participants or, in the case of minors, their legal guardian or next of kin. In addition, the University of Washington Human Subjects Division approved an earlier version of this study (STUDY00002855) that did not include subjects under the age of ten.

## Consent for Publication

No applicable.

## Author’s contributions

YS designed the study. XX and EC led the data analysis and wrote the original draft. DW acquired the funding. XX, EC, DW, CL and YS made important contributions to the revision of the manuscript. JA, RK, MN, EN, HN, EO, JP and GC participated in the revision of the manuscript. All authors have read and approved the final manuscript.

